# Ecologic correlation between underlying population level morbidities and COVID-19 case fatality rate among countries infected with SARS-CoV-2

**DOI:** 10.1101/2020.04.28.20082370

**Authors:** Evaezi Okpokoro, Victoria Igbinomwanhia, Elima Jedy-Agba, Gbenga Kayode, Ezenwa James Onyemata, Alash’le Abimiku

**Affiliations:** International Research Center of Excellence, Institute of Human Virology Nigeria; Vaccine for Africa University of Cape Town; University of Maryland, Baltimore

## Abstract

**Background:** The ongoing Coronavirus disease 2019 (COVID-19) pandemic is unprecedented in scope. High income countries (HIC) seemingly account for the majority of the mortalities considering that these countries have screened more persons. Low middle income countries (LMIC) countries may experience far worse mortalities considering the existence of a weaker health care system and the several underlying population level morbidities. As a result, it becomes imperative to understand the ecological correlation between critical underlying population level morbidities and COVID-19 case fatality rates (CFR).

**Method:** This is an ecological study using data on COVID-19 cases, prevalence of COPD, prevalence of tobacco use, adult HIV prevalence, quality of air and life expectancy. We plotted a histogram, performed the Shapiro-Wilk normality test and used spearman correlation to assess the degree of correlation between COVID-19 case fatality rate (CFR) and other covariates mentioned above.

**Result:** As at the 31^st^ of March 2020, there were a total of 846,281 cases of COVID-19 from 204 countries and a global case fatality rate of 5% (range 0% to 29%). Angola and Sudan both had the highest CFR of 29%, while Italy had the highest number of deaths (i.e. 12,428) as at 31^st^ of March 2020. Adult HIV prevalence has a significant but weak negative correlation with CFR (correlation coefficient = - 0.24, p value =0.01) while all the other variables have positive correlation with CFR due to COVID-19 though not statistically significant. Of the 204 countries analyzed, only 11 countries (i.e. 5%) had complete datasets across all 5 population level morbidities (i.e. prevalence of COPD, prevalence of tobacco use, life expectancy, quality of air, and adult HIV prevalence variables). Correlations of CFR from these 11 countries were similar to that from the 204 countries except for the correlation with quality of air and prevalence of tobacco use. *Conclusion:* While we interpret our data with caution given the fact that this is an ecological study, our findings suggest that population level factors such as prevalence of COPD, prevalence of tobacco use, life expectancy and quality of air are positively correlated with CFR from COVID-19 but, adult HIV prevalence has a weak and negative correlation with COVID-19 CFR and would require extensive research.

## Background

The ongoing Corona virus disease 2019 (COVID-19) pandemic is unprecedented in scope. As at the 31st March 2020, 846,281 persons from 204 counties have been infected. Of these numbers, 31,599 were seriously or critical ill and 41,482 had died.^1^ High-income (HIC) countries seemingly account for the majority of the mortalities worldwide. Incidentally these countries have screened a larger proportion of their populations’ for COVID-19. Despite the presence of efficient health care systems in these countries, they have been unable curb the persistent rise in mortalities. This scenario suggests that should there be an exponential rise in COVID-19 cases among low and middle income countries (LMICs), these countries may experience far worse mortalities considering the existence of a weaker health care system and the several underlying population level morbidities. As a result, it becomes imperative to understand the ecological relationship between critical underlying population level morbidities globally and mortalities associated with SAR-CoV-2 infection.

According to anecdotal evidence, most mortalities following COVID-19 occurs among the aging populations but to our knowledge, there has been no documented evidence to confirm this. In addition, considering the high burden of HIV in LMICs particularly in sub-Saharan Africa (SSA), there are serious concerns of the impact of HIV on mortalities associated with COVID-19. COVID-19 is caused by an RNA virus (i.e. SAR-CoV-2) which affects mainly the respiratory system, therefore, countries with a high burden of underlying respiratory diseases such as chronic obstructive pulmonary disease (COPD) may experience higher mortalities compared to those with lower prevalence of respiratory diseases. Factors such as cigarette smoking and quality of air may impact on the respiratory system leading to higher mortalities due to COVID-19 in such countries. Generating quick preliminary evidence through an ecologic study could provide vital insight at the population level, to understanding factors correlated with COVID-19 case fatality rates especially as the incidence rapidly increases in low and middle income countries.

## Methods

This is an ecological study using data from several open sources on the internet and from systematic reviews. We obtained data on COVID-19 cases and mortality from the worldometers as at 31^st^ March 2020 and life expectancy data of 2016 from online sources^2^. We also obtained data on prevalence of COPD, adult HIV, tobacco use and quality of air from other online sources and journals^3–6^. We plotted a histogram to assess the distribution of the data as well as performed the Shapiro-Wilk normality test. Thereafter, we provided appropriate summary statistics. We also performed two-way scatterplots involving key variable and thereafter used a spearman correlation test to assess the degree of correlation between COVID-19 case fatalities rates and other covariates (table 2).

## Results

In our study, we define case fatality rate (CFR) as the number of deaths due to COVID 19 divided by the number of confirmed cases officially reported. As shown in table 1, a total of 204 countries were identified with cases of COVID-19 as at 31^st^ March 2020. The mean number of deaths per day was 512, while the mean number of new cases was 10,448. The global case fatality rate was 5% while it ranges from 0% to 29%. Angola and Sudan had the highest CFR while Italy had the highest number of deaths (i.e. 12,428). Of the 204 countries infected with COVID-19, 33 (16%) had mortality rates greater than 5% (i.e. above the global average).

**Table 1:**
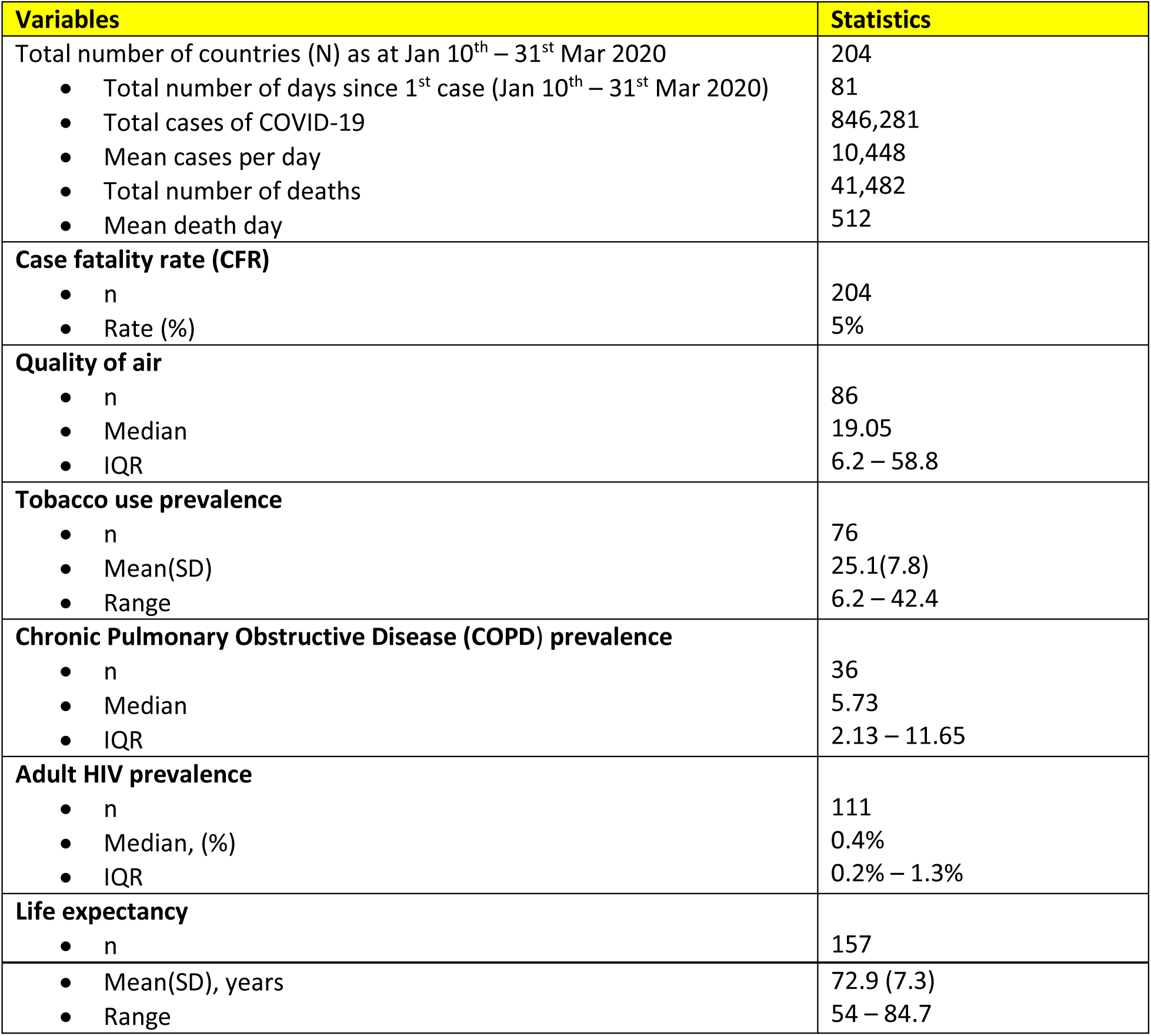
Descriptive statistics.

As shown in table 2 and likewise in the scatterplot (fig 1), adult HIV prevalence has a significant weak and negative correlation with case fatality rate associated with COVID-19 while all the other variables have positive correlation with COVID-19 though not statistically significant.

**Table 2:** Spearman correlation of underlying population level morbidity and CFR due to COVID-19.

| Variable | N | Correlation coefficient (Rho) | p-value | Comment |
| --- | --- | --- | --- | --- |
| COPD prevalence | 36 | 0.28 | 0.09 | Weak and positive correlation |
| Tobacco use prevalence | 76 | 0.01 | 0.91 | Very weak and positive correlation |
| Life expectancy | 157 | 0.09 | 0.24 | Very weak and positive correlation |
| Quality of air | 86 | 0.02 | 0.84 | Very weak and positive correlation |
| Adult HIV prevalence | 111 | -0.24 | **0.01** | Weak and negative correlation |

\*.00-.19 "very weak", .20-.39 "weak", .40-.59 "moderate", .60-.79 "strong", .80-1.0 "very strong"

**Figure 1:**
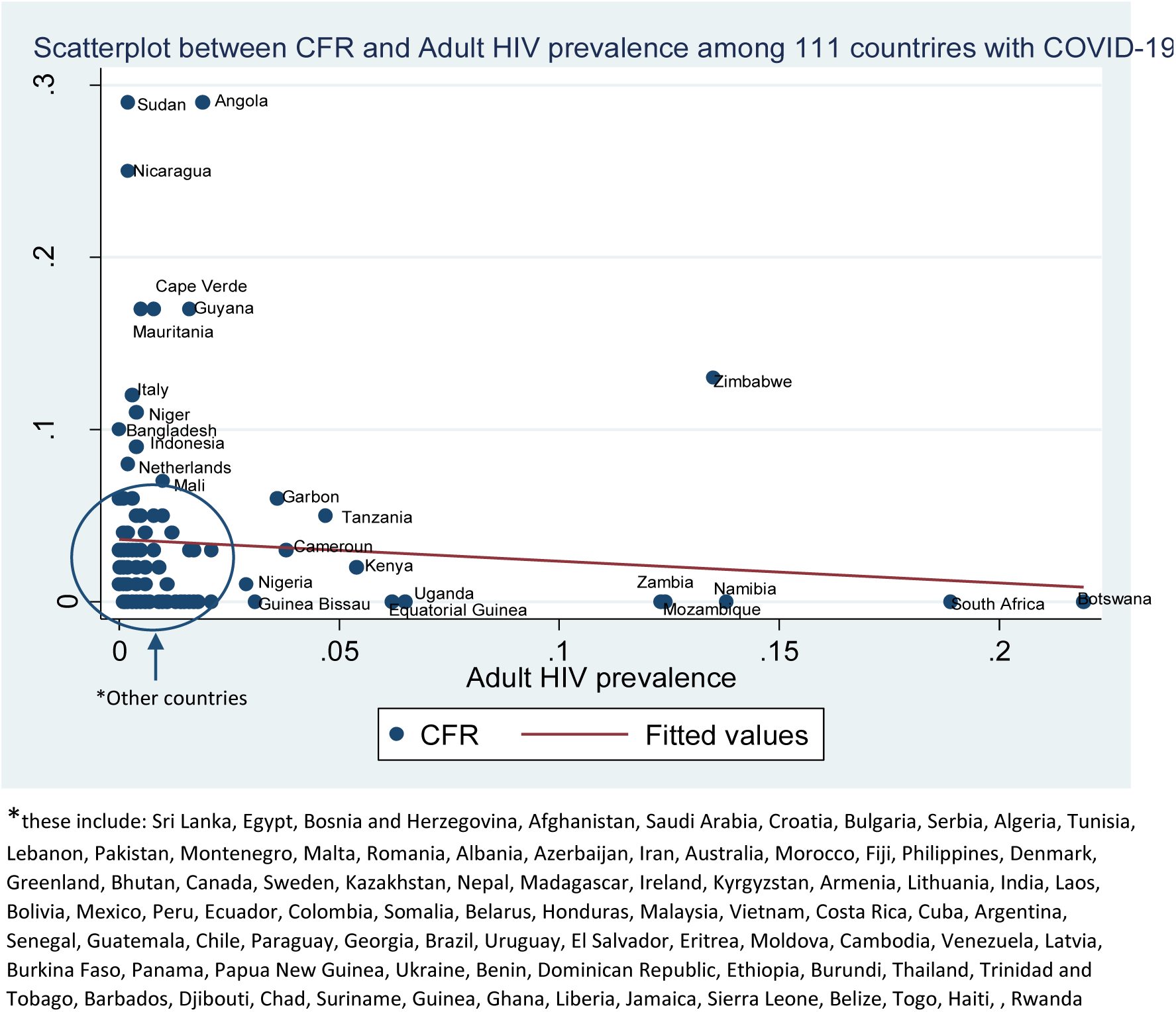
Correlation scatterplot of CFR vs Adult HIV prevalence (among 111 countries) as at 31^st^ March 2020

Of the 204 countries, only 11 countries (i.e. 5%) had complete datasets covering all 5 population level morbidities (i.e. COPD prevalence, tobacco use prevalence, life expectancy, quality of air and adult HIV prevalence variables). Our findings from analyzing dataset of these 11 countries (table 3) were similar with the 204 countries (table 2) except for the correlation between quality of air, tobacco use and CFR due to COVID-19.

**Table 3:** Spearman correlation between CFR and population level morbidities among ONLY 11 out of 204 countries with complete dataset of all 5 co-variates.

| Variable | N | Correlation coefficient (Rho) | p-value | Comment* |
| --- | --- | --- | --- | --- |
| COPD prevalence | 11 | 0.62 | **0.03** | Strong and positive correlation |
| Tobacco use prevalence | 11 | -0.51 | 0.1 | Moderate and negative correlation |
| Life expectancy | 11 | 0.9 | **0.00** | Very strong and positive correlation |
| Quality of air | 11 | -0.72 | **0.01** | Strong and negative correlation |
| Adult HIV prevalence | 11 | -0.25 | 0.45 | Weak and negative correlation |

\*.00-.19 "very weak", .20-.39 "weak", .40-.59 "moderate", .60-.79 "strong", .80-1.0 "very strong"

## Discussion

This ecological study has demonstrated that a substantial amount of the variation observed in case fatality rate associated with COVID-19 across 204 countries as at 31^st^ March 2020 could be explained by five population level morbidities such as; prevalence of COPD, prevalence of tobacco use, adult HIV prevalence, quality of air and life expectancy. Our findings suggest that CFR was highest in Angola and Sudan as at 31^st^ March 2020. We found varying levels of positive correlations between COPD prevalence, life expectancy and quality of air with CFR in COVID-19 disease.

Our findings suggest that adult HIV prevalence is seemingly protective. However, we must interpret this finding with caution given the weak negative correlation with CFR in COVID 19 (correlation coefficient= - 0.24, p=0.01). This negative relationship was maintained even when we had a closer analysis involving a subset of only 11 out of 204 countries which had complete dataset across the 5 variable (see fig 2). Thus countries with higher prevalence of HIV may experience lower mortalities compared with those with lower HIV prevalence. This was a rather surprising finding as we expected that populations with higher adult HIV prevalence would have more immunocompromised persons and therefore have higher COVID-19 associated CFR. In accordance with our findings, some experts have suggested the possibility of using antiretroviral therapies such as lopinavir/ritonavir against COVID 19 and clinical trials involving antiviral medications are currently ongoing^7^. However, more extensive research studies are needed in this area in order to confirm or refute this finding. In one clinical trial involving severely ill COVID-19 patients, there was no significant effect of lopinavir–ritonavir treatment in terms of reducing mortality, increasing clinical improvement and decreasing throat viral RNA detectability among these patients. Thus, considering the combination of lopinavir–ritonavir with other antiviral agents to boost its effects might be an option in future studies ^8^

**Figure 2.**
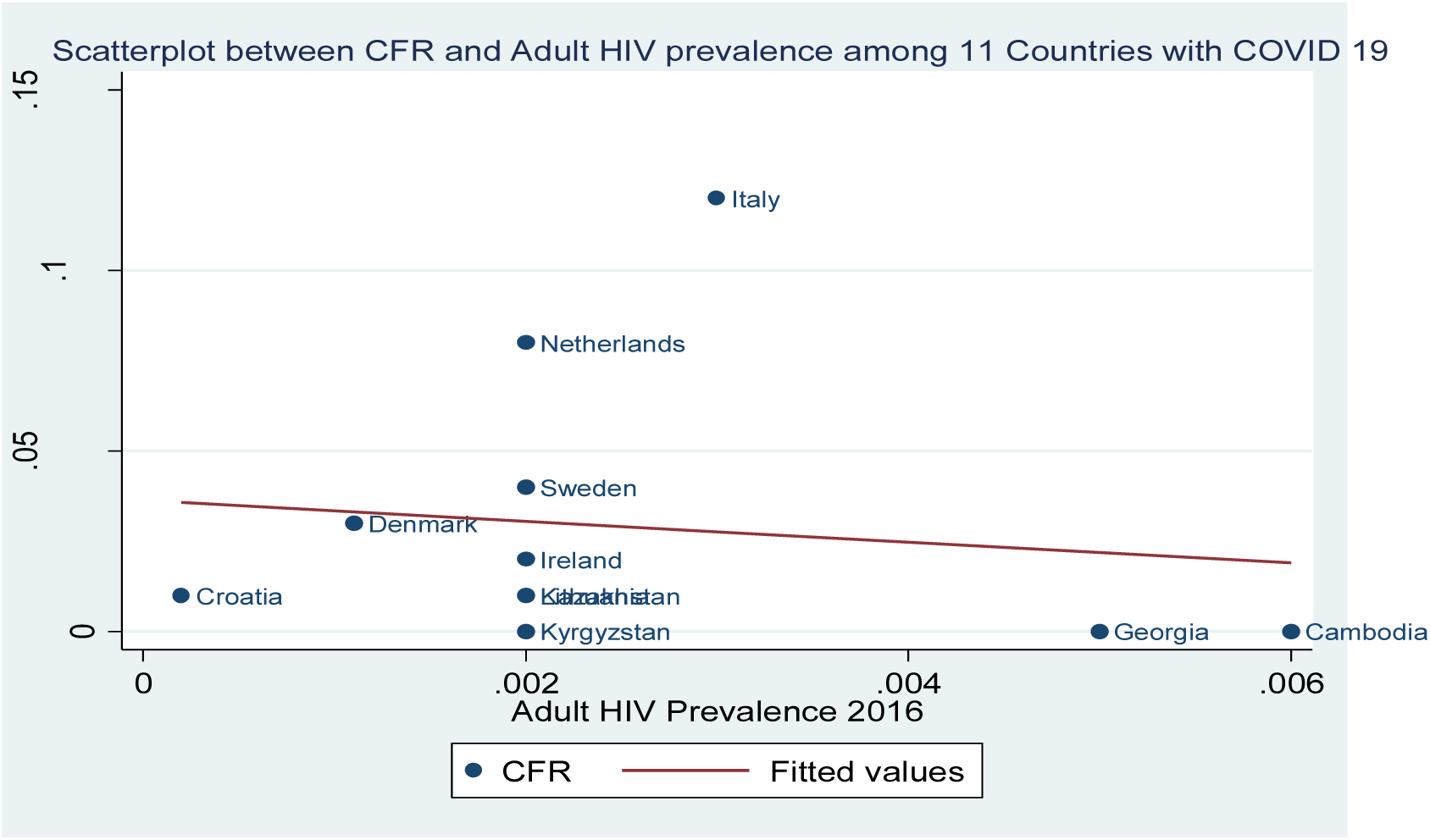
Correlation scatterplot of CFR vs Adult HIV prevalence among 11 Countries only as at 31^st^ March 2020

We found life expectancy to have a very weak but positive correlation with CFR (correlation coefficient 0.09; p=0.24). However, this correlation was strongly positive when we reviewed a subset of 11 countries (see fig 3). Thus countries with higher life expectancy were likely to experience a higher CFR when compared with countries with lower life expectancy. Similar demographics or findings have been informally reported by researchers in other countries. In South Africa, 10 out 18 (55%) mortality occurred among those 70 years and over while the average age was 68 years^9^. Similarly, in Italy, about 63.3% of mortality were among those over 70years and while in Spain, a rate of 23% was reported among in those over 90 years of age^10,11^.

**Figure 3:**
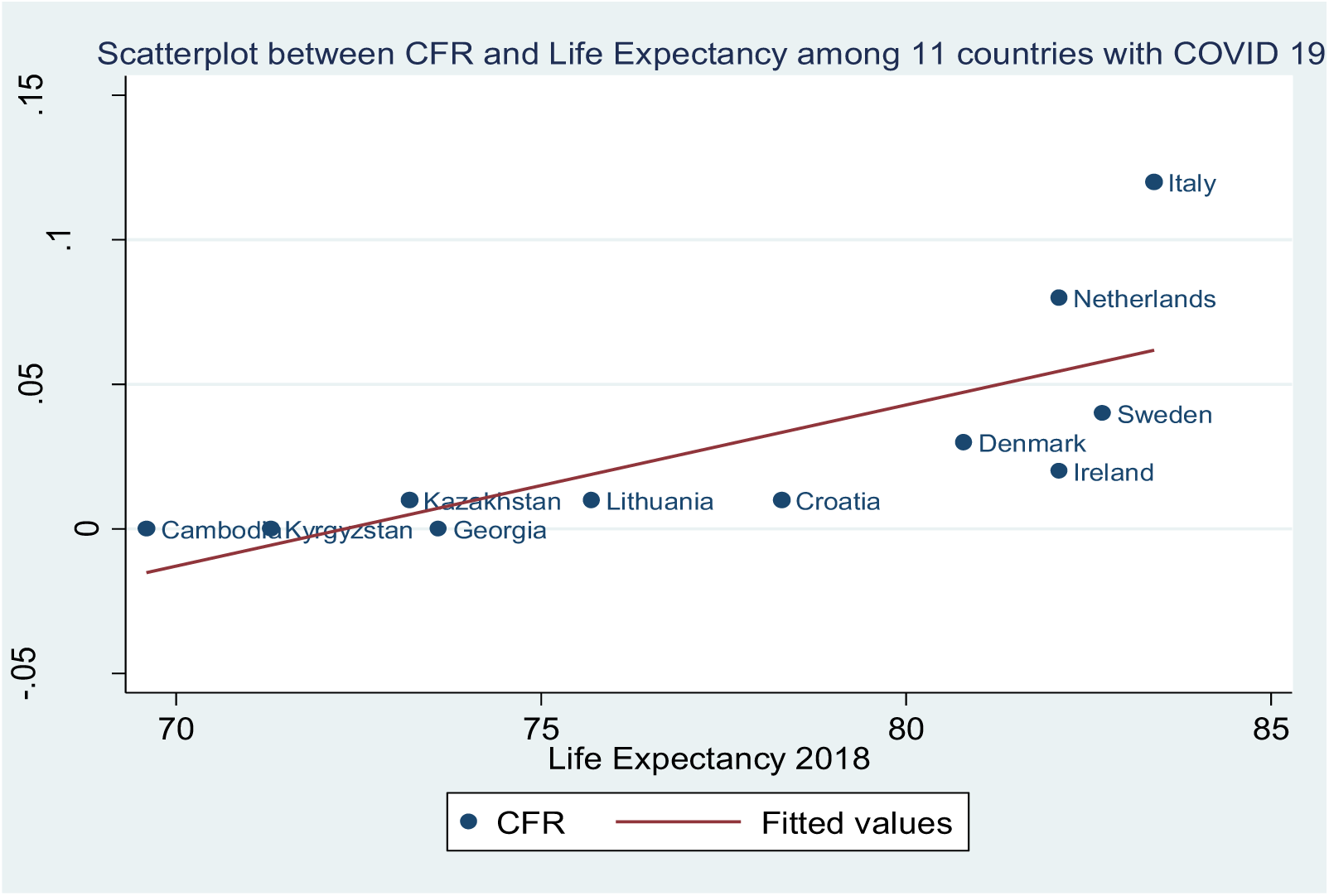
Correlation scatterplot-CFR vs Life Expectancy as at 31^st^ March 2020

Considering the fact that COVID 19 affects the respiratory system cases, we found a weak but positive correlation between the prevalence of COPD and CFR due to COVID-19 (correlation coefficient = 0.09, p value =0.09). This correlation was stronger when we reviewed a subset of only 11 countries with complete data out 204 (see fig 4). This relationship may be due to the underlying relationship between COPD and older populations^12^. Thus, countries with older populations or higher life expectancy would likely have a higher prevalence of COPD which in turn could lead to higher CFR due to COVID-19^13^.

**Figure 4:**
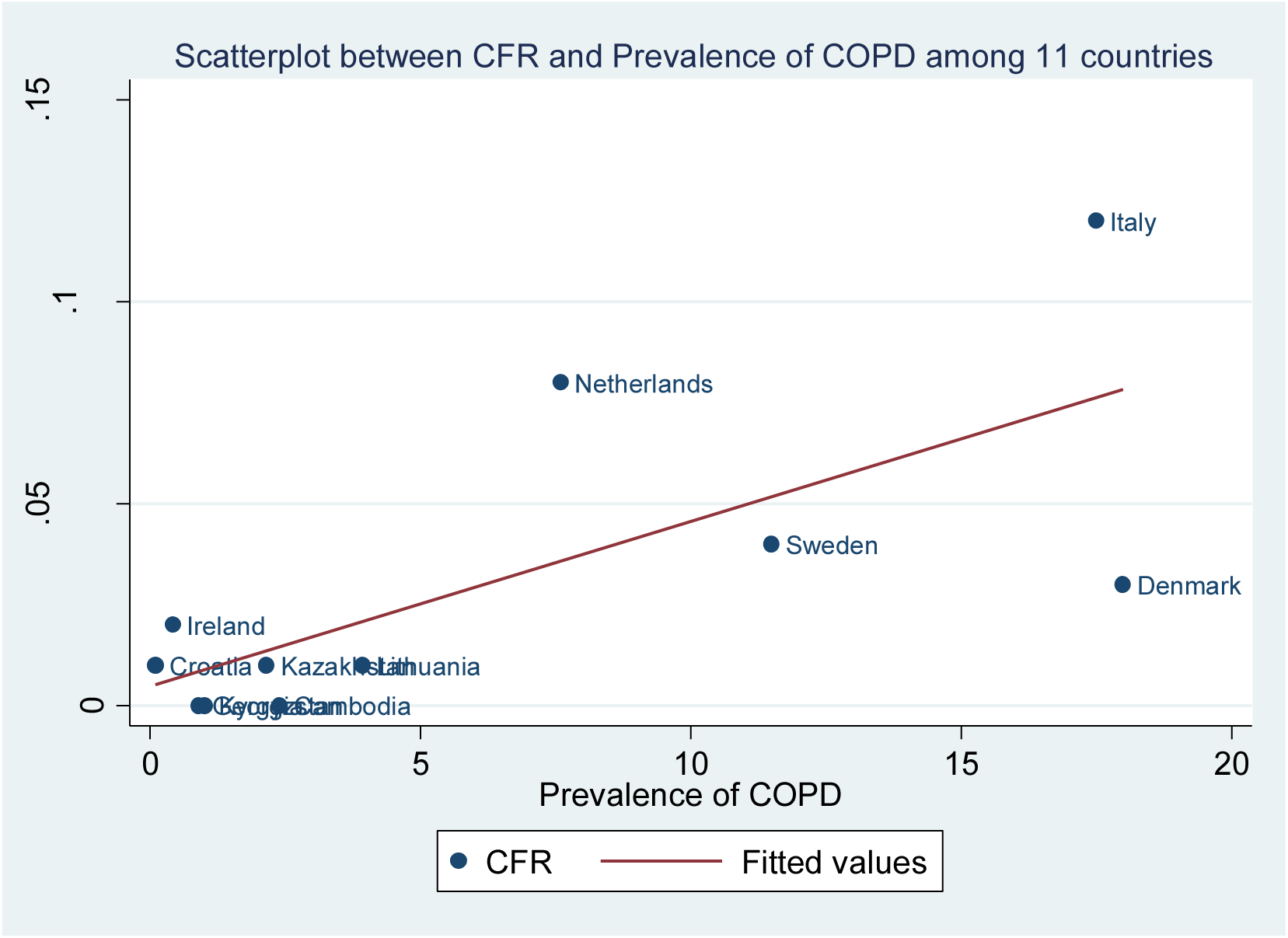
Scatterplot showing Correlation between COPD and CFR in COVID-19 as at March 2020.

Our findings on the relationship between CFR due to COVID-19 and prevalence of tobacco use and/or quality of air, showed that both factors were very weakly and positively correlated to CFR (correlation coefficient= 0.01 and 0.02 respectively). However, this finding was not statistically significant (pvalue =0.91 and 0.84 respectively). Thus increased tobacco consumption and higher ranking on quality of air (higher ranking meaning higher air pollution) is correlated with higher CFR due to COVID-19. Previous studies have reported the link between respiratory viral infection and air pollutions. According to these studies, respiratory viral infection increased with air pollution^14^. Also a recent study by researchers at Harvard University reports a higher CFR due to COVID-19 among counties within the US with higher air pollution^15^. Surprisingly, when we reviewed data of a subset of only 11 out of 204 countries with complete datasets, there was a reversal from positive to negative correlation between CFR and the prevalence of tobacco use and/or quality of air. We noticed a negative and strong correlation between CFR due to COVID-19 (correlation coefficient = −0.51 and −0.72 respectively), though not statistically significant. This may be due to the small sample size but it is worthy of future research.

Countries in LMICs are increasing the number of persons being tested for SARS-CoV-2 infection through innovative new technologies for SARS-COV2 testing. Despite the rise in tests being conducted, it is likely that these countries may not experience the high mortality due to COVID 19 as is currently being reported from HIC countries due to many reasons. Firstly, owing to the few tests currently being done, the number likely to die from a confirmed diagnosis of COVID-19 would be underestimated. Secondly, the earlier awareness and lead time in preparations by LMICs prior to the advent of COVID-19 might reduce the spread and impact. More so, findings from our study suggest that a higher prevalence of adult HIV as experienced in LMIC might be protective against higher CFR due to COVID-19. Lastly, and hopefully, ongoing clinical trials might be successful before LMIC begin to experience such a comparable high mortality.

An important limitation of this study is the ecological nature of the study, hence, findings must be interpreted with caution and should not be directly extrapolated to individual levels. Secondly, COVID-19 is a rapidly evolving disease and total numbers of confirmed cases and mortality figures are constantly changing. Thirdly, the datasets used for our analysis were publicly available on the internet and may not be accurate as officially published estimates by the respective countries included in our study. However, our study presents data from 204 countries worldwide on 846, 281 confirmed cases of COVID-19. Additionally, this study also provides useful information on population level factors that may contribute to the noticeable differences in mortality rates worldwide, which is an issue of significant interest to epidemiologists, clinicians, and public health policy makers worldwide.

## Conclusion

Our findings suggest that population level factors such as COPD prevalence, prevalence of tobacco use, life expectancy and quality of air are positively correlated with CFR from COVID-19. While adult HIV prevalence has a weak and negative correlation with COVID-19 CFR. Extensive research is required to investigate these population level factors at the individual level in order to provide information that can be generalizable.

## Data Availability

Publicly available on request

